# Have infection control and prevention measures resulted in any adverse outcomes for care home and domiciliary care residents and staff?

**DOI:** 10.1101/2022.05.04.22274657

**Authors:** Review Team, Llinos Haf Spencer, Ned Hartfiel, Annie Hendry, Bethany Anthony, Abraham Makanjuola, Report Team, Nathan Bray, Dyfrig Hughes, Clare Wilkinson, Deb Fitzsimmons, Rhiannon Tudor Edwards

**Affiliations:** Bangor University; Swansea University

## Abstract

**What is a Rapid Review?:** Our rapid reviews use a variation of the systematic review approach, abbreviating or omitting some components to generate the evidence to inform stakeholders promptly whilst maintaining attention to bias. They follow the methodological recommendations and minimum standards for conducting and reporting rapid reviews, including a structured protocol, systematic search, screening, data extraction, critical appraisal and evidence synthesis to answer a specific question and identify key research gaps. They take 1-2 months, depending on the breadth and complexity of the research topic/question(s), the extent of the evidence base and type of analysis required for synthesis.

**Background / Aim of Rapid Review:** Care for older and vulnerable people must sustain core infection prevention and control (IPC) practices and remain vigilant for COVID-19 transmission to prevent virus spread and protect residents and healthcare professionals from severe infections, hospitalisations and death.

However, these measures could **potentially lead to adverse outcomes** such as decreased mental wellbeing in patients and staff. A recent publication by Public Health England examines the effectiveness of IPC practices for reducing COVID-19 transmission in care homes (Duval et al., 2021). **We explore evidence relating to adverse outcomes from IPC practices to help inform policy recommendations and identify gaps within the literature where further research can be prioritised**.

**Key Findings:** *Extent of the evidence base:*

- 15 studies were identified: 14 primary studies and one rapid review

*Recency of the evidence base:*

- Of the primary studies, six were published in 2020 and eight were published in 2021
- The rapid review was published in 2021.

*Summary of findings:* **This rapid review focuses on adverse outcomes resulting from increased IPC measures** put in place during the COVID-19 pandemic. Whilst there is some evidence to show that there may be a link between IPC measures and adverse outcomes, causation cannot be assumed. - During the COVID-19 restrictions, **the cognition, mental wellbeing and behaviour of residents in care homes were negatively affected**
- Increased IPC procedures during the COVID-19 pandemic **increased stress and burden among care staff** because of increased workload and dilemmas between adhering well to IPC procedures and providing the best care for the care recipients
- COVID-19 IPC procedures were not well developed at the beginning of the COVID-19 pandemic, but evidence from 2021 suggests that **good adherence to IPC measures can enable visitations by family members and medical professionals into care homes**
- **Only one study investigating domiciliary care was found**. Therefore, it is difficult to make conclusions related specifically to this care setting
- **No published studies have reported on the costs or cost-effectiveness of IPC measures** or have explored the cost implications of adverse outcomes associated with IPC measures

*Best quality evidence:* Only one study was deemed as high quality based on the quality appraisal checklist ranking. This was a mixed methods study design (Tulloch et al., 2021).

**Policy Implications:** Since March 2020, there have been many changes to government guidelines relating to procedures to keep the population safe from COVID-19 harm. Policies vary according to country, even within the UK. Important issues such as care home visitation policies have changed in such a way that care home staff have felt it difficult to keep up with the changes, which in itself increased the burden on those staff. The following implications were identified from this work:

- IPC policies should be clear, concise and tailored to care homes and domiciliary care settings
- Increased attention to workforce planning is needed to ensure adequate staffing and to reduce individual burden
- Restrictions (e.g. visitation) for care home residents needs to be balanced by additional psychological support
- Further research with robust methods in this area is urgently needed especially in the domiciliary care setting

**Strength of Evidence:** One limitation is the **lack of high-quality evidence from the included studies**. Confidence in the strength of evidence about adverse outcomes of COVID-19 IPC procedures was rated as ‘low’ overall. Whilst the majority of studies achieved a ‘moderate’ score based on the quality appraisal tools used, due to the nature of the methods used, the overall quality of evidence is low.

## 1. BACKGROUND

This Rapid Review is being conducted as part of the Wales COVID-19 Evidence Centre Work Programme. The above question was suggested by the Technical Advisory Group (TAG) Policy Modelling Subgroup.

SARS-CoV-2 is a very infectious coronavirus with transmission primarily through respiratory particles, airborne transmission and via fomite contact (Social Care Institute for Excellence, 2021; Vardoulakis, Sheel, Lal, & Gray, 2020). According to itccovid.org, at the beginning of the COVID-19 pandemic, there were 18,075 adult care homes in total across the UK with around 500,598 adult care home residents. Of these, 425,408 were in England, 14,935 in Northern Ireland, 35,989 in Scotland and 23,766 in Wales (Bell et al., 2020).

Adults living in care home settings or receiving domiciliary care are at high risk of being affected by COVID-19 (Lauter et al., 2021; Ouslander & Grabowski, 2020). Care homes providing services to older people with dementia recorded most deaths from COVID-19 (Morciano, Stokes, Kontopantelis, Hall, & Turner, 2021). Given the difficulties complying with physical distancing, masking and hand hygiene with such a vulnerable population, this is unsurprising. However, the risk of SARS-CoV-2 entering nursing care establishments compared with residential care homes was lower. This might be because of the protective effect of staff with nursing backgrounds and infection, prevention and control (IPC) training (Morciano et al., 2021).

Since the beginning of the COVID-19 pandemic in March 2020 until 2^nd^ April 2021, there were 173,974 deaths of care home residents in England and Wales. This is an increase of 19.5% compared with the five-year average (145,560 deaths). Of these, 42,341 involved COVID-19 (24.3% of all deaths of care home residents) (Office for National Statistics, 2021a).

Age, gender, ethnicity, deprivation and physical disability were also risk factors for dying from COVID-19 (Niedzwiedz et al., 2020). Public Health England (Public Health England, 2020) stated that among people already diagnosed with COVID-19, people aged 80 or older were seventy times more likely to die than those under 40 years of age. The risk of death among those diagnosed with COVID-19 was also higher in males, in those living in more deprived areas, and in Black, Asian and Minority Ethnic (BAME) groups (Public Health England, 2020). People with learning disabilities were also at a higher risk of contracting and dying from COVID-19 (Kavanagh et al., 2021). With regards to people with physical disabilities, many were significantly affected by the national lockdowns and restrictions. The risk of death from COVID-19 was 3.1 times greater for men with disabilities and 3.5 times greater for women with disabilities than for men and women without disabilities (Office for National Statistics, 2021b). At the start of the pandemic, there were many COVID-19 related publications, but only 11 papers highlighting issues for people living with disabilities, according to a 2021 rapid review (Lebrasseur et al., 2021).

There was a lack of sufficient COVID-19 testing and a lack of personal protective equipment (PPE) at the beginning of the pandemic in the UK and worldwide (Lipsitz et al., 2020; Telford et al., 2020). A lack of access to adequate PPE and COVID-19 testing in care homes posed a major risk of COVID-19 spreading. Care homes were identified in March 2020 as one of the most significant potential risks for COVID-19 spreading due to the number of elderly residents with underlying health conditions (Iacobucci, 2020). The transfer of patients from hospitals to homes without adequate testing also led to the increased spreading of the disease (House of Commons, Health and Social Care, Technology, & Committees, 2021).

Since July 2021, the Department of Health and Social Care outlined two main objectives regarding infection prevention and control in adult social care. Firstly, to reduce the rate of COVID-19 transmission within and between care settings through effective IPC practices and increase uptake of staff vaccination. Secondly, to conduct testing of staff and visitors in care homes and high risk supported living and extra care settings, to enable close contact visiting where possible (Department of Health & Social Care, 2021). A strong IPC program is critical to protect residents, care workers and healthcare personnel (HCP) (Bell et al., 2020; Shah et al., 2021). According to the NHS Confederation report in September 2021, acute NHS Trusts are currently spending around £400k per month over and above their budget for increased IPC measures, e.g. cleaning, site security and waste disposal (NHS Confederation, 2021).

By October 2021, with an increase in COVID-19 lateral flow testing and PCR testing, and an improved NHS Test and Trace system aligned with the success of COVID-19 vaccination programmes, care homes and domiciliary care services began to resume normal practices, and restrictions started to relax. Nevertheless, care for older adults must sustain core IPC practices and remain vigilant for COVID-19 transmission to prevent spread and protect residents and healthcare professionals from severe infections, hospitalizations, and death (Low et al., 2021). The UK and Welsh Governments are providing guidance on IPC in care homes as well as guidance on visits to care homes (NHS National Services Scotland, 2021; Public Health England, 2021a; UK Government, 2021; Welsh Government, 2021).

Although core IPC measures, such as more frequent cleaning and changing PPE, are essential for reducing virus transmission, evidence indicates that these measures may have generated some adverse outcomes such as decreased mental wellbeing in patients and staff. Additional burden experienced by caregivers can lead to decreased staff morale and burnout (House of Commons Committees, 2021; Royal College of Nursing Wales, 2021), which leads to increased staff absenteeism (Bell et al., 2020; Groenewold et al., 2020).

There has been a loss of care staff due to Brexit and staff shortages due to healthcare workers seeking new employment (Dalingwater, 2021). Loss of care staff can be attributed to COVID-19 vaccine hesitancy, such as fear of the unknown effect of a COVID-19 vaccine on future pregnancies (Woolf et al., 2021). Workforce planning to ensure adequate health and social care staff is urgent in the UK (House of Commons, 2021). The Royal College of Nursing Wales have stated that close integration of health and social care means that nursing shortages in the care home sector and/or poor workforce planning are a significant risk for the NHS (Royal College of Nursing Wales, 2020, 2021).

This review complements previous work by Public Health England (Public Health England, 2021b), which explored the effectiveness of IPC measures in reducing transmission of COVID-19 in care homes. In our rapid review, we did not include transmission, hospitalisation or mortality rates due to the comprehensive nature of the Public Health England report, which covered these areas within the same time frame (Duval et al., 2021).

IPC procedures refer to a range of measures used by care staff to reduce the risk of COVID-19 transmission through methods such as the use of face masks and other personal protective equipment (PPE), additional cleaning protocols, routine COVID-19 testing, social distancing, hand washing, isolation, changes to visitation, increased ventilation and other measures.

### 1.1 Review question

Have infection control and prevention measures resulted in any adverse outcomes for care home and domiciliary care residents and staff?

### 1.3 Objectives of this rapid review

The rapid review methods, including quality appraisal methods, are presented in full in Section 4 of this report. However, our review focuses on a variety of adverse outcomes relating to IPC procedures across the different stakeholder groups, such as:

- Recipients of care: wellbeing and quality of life
- Care staff: buy-in and support for IPC procedures, acceptability/adherence to IPC procedures, mitigation of fatigue towards IPC procedures; and wellbeing/quality of life outcomes
- Costs: ongoing costs, cost-effectiveness and potential savings.

## 2. RESULTS

### 2.1 Overview of the Evidence Base

In this rapid review, 15 papers were identified for inclusion that reported on adverse outcomes related to IPC procedures during the COVID-19 pandemic; 14 were primary studies, and one was a rapid review of the evidence within a one-year period.

A narrative summary of the 15 studies is provided below. The rapid review findings are categorised into three main themes: resident outcomes, staff outcomes and COVID-19 guidance and context. Quality appraisals for each of the included studies can be found in Appendix 1.

### 2.2 Adverse outcomes

#### 2.2.1 Resident outcomes

##### Mental health of care home residents

IPC measures designed to limit social contact such as reduced visitation and social distancing were reported to be associated with adverse mental health outcomes in seven included studies. A cohort study by El Haj et al., (2020) highlighted the increase in depression and anxiety in a population of 58 care home residents with Alzheimer’s disease in France. Participants reported higher levels of depression whilst isolation measures were in place (M = Mean, SD = standard deviation) (M = 14.21, SD = 3.17) than before (M = 12.34, SD = 4.10) the COVID-19 crisis (Cohen’s d = 0.80, Z = -2.84, p = .005). Participants also reported higher levels of anxiety during (M = 13.24, SD = 3.39) than before (M = 11.38, SD = 4.36) the COVID-19 crisis (Cohen’s d = 0.81, Z = -2.86, p = .004) (El Haj, Altintasb, Chapelete, Kapogiannisg, & Galloujb, 2020).

Mental health status was also measured by Van der Roest et al., (2020) in a cross-sectional survey. Mood in nursing home residents in the Netherlands was assessed with the Mental Health Inventory 5-Index (MHI-5; range 0-100, scores <60 indicate poor mental health). Mean MHI-5 score for residents was 56.6 (SD 20.4). 51% had scores <60. Only 27% of relatives reported no change in residents’ mood status. Happiness was reported less often, and sadness was more often reported by families of residents without cognitive impairment than with cognitive impairment (P < .000; P < .008, respectively). Loneliness was reported by 77% of residents. Staff classified residents without cognitive impairment as more lonely than residents with cognitive impairment (P < .006) (Van der Roest et al., 2020).

In a qualitative exploratory study, Sizoo et al., (2020) found that at the beginning of the COVID-19 pandemic (April-May 2020) elderly care practitioners observed loneliness, depressive symptoms, decreased intake, increased somatic symptoms (i.e. pain), physical deterioration and, in psychogeriatric residents’, rapid cognitive decline and changes in neuropsychiatric symptoms including agitation and aggression as a result of the restrictions in visitations. In the study of residents in a Canadian care home by Iaboni et al., (2020) reduced visitation and isolation measures were associated with increased levels of agitation and aggression resulting in increased drug prescriptions.

A Belgian qualitative study by Kaelen et al., (2021) found that nursing home residents experienced losses of freedom, social life, autonomy, and recreational activities that deprived them of their basic psychological needs following the introduction of IPC measures such as isolation, social distancing and reduced or paused visitation. These losses of freedom impacted on mental wellbeing, and residents expressed feelings of depression, anxiety, frustration and decreased meaning and quality of life (Kaelen et al., 2021). In the Schweighart et al., (2021) qualitative study, nursing home residents in Germany also reported increased boredom and inactivity due to the COVID-19 restrictions on social interaction, mask-wearing and ventilation measures. The residents in this study complained of feeling cold due to the ventilation changes, having limited contact with other residents in the home and felt that wearing masks was a nuisance, because they impeded breathing and made wearing glasses and hearing aids problematic (Schweighart, Klemmt, Neuderth, & Teti, 2021).

The rapid review conducted by Suarez-Gonzalez et al., (2021) aimed to examine and summarise the global research evidence describing the effect of COVID-19 isolation measures on the health of people living with dementia worldwide. IPC measures to limit social contact, such as lockdowns and confinement measures brought about by the COVID-19 pandemic, have reportedly damaged the cognitive and psychological health and functional abilities of people with dementia globally (Suárez-González, Rajagopalan, Livingston, & Alladi, 2021). The authors of this review found 15 eligible papers, examining a total of 6,442 people with dementia. Of these papers,13 focused on people living in the community and two in care homes. It may be that not all of these studies are directly relevant to this review, as not all dementia patients living in the community will receive domiciliary care. 60% (9/15) of the studies reported changes in cognition, with 77% (7/9) of them describing declined cognition in >50% of respondents. 93% (14/15) of the studies reported worsening or new onset of behavioural and psychological symptoms. 46% (7/15) of the studies reported changes in daily function, and six papers reported a functional decline in a variable proportion of the population studied.

The authors of this review (Suarez-Gonzalez et al., 2021) state the need for four specific calls for action:

- Formal and informal carers should be prioritised for vaccination
- Remote working for informal carers should be extended beyond the pandemic
- Social contact outdoors with appropriate PPE to be used to facilitate social activity
- In countries where vaccination rates are poorer, measures should be taken to facilitate safe contact

The searches for this review were conducted up to February 2021, and at that time, very little information was available about the impact of COVID-19 on people residing in care homes.

**Bottom line. During the COVID-19 restrictions the cognition, mental wellbeing and behaviour of residents in care homes was negatively affected**.

#### 2.2.2 Care staff outcomes

##### Wellbeing of care home staff

The qualitative study by Kaelen et al., (2021) found that nursing home staff in Belgium felt unprepared for the challenges posed by the COVID-19 pandemic. Due to the lack of pandemic guidelines, PPE availability and clarity around the organisation of care at the start of the pandemic, nursing home staff were confronted with professional and ethical dilemmas and felt ‘trapped’ between adhering to IPC measures and maintaining residents’ wellbeing. They witnessed the detrimental effects of the measures imposed on their residents (Kaelen et al., 2021).

##### Work related stress

A cross-sectional survey by Nestor et al., (2021) found that nursing and healthcare staff working exclusively in elderly care settings reported stress and anxiety due to constantly changing IPC protocols (specifically related to PPE) with little to no explanation of the rationale driving the protocol changes. Constantly changing IPC protocols was ranked 4^th^ highest of 17 workplace stressors by nursing and healthcare staff (Nestor, O’ Tuathaigh, & O’ Brien, 2021). In the Sizoo et al., (2020) qualitative study, care home staff stated that there were moral issues with adhering to IPC measures. Staff described experiencing emotions of guilt and injustice when implementing IPC measures to reduce social contact, such as restricted visitation (Sizoo, Monnier, Bloemen, Hertogh, & Smalbrugge, 2020). Iaboni et al., (2020) also found that IPC measures requiring staff to frequently change PPE resulted in increased pressure and stress levels when providing care (Iaboni et al., 2020).

##### Burden of IPC measures

In the rapid review by Suarez-Gonzales et al., (2021) 93% (14/15) of studies reported worsening or new onset of behavioural and psychological symptoms in people living with dementia. 46% (7/15) studies reported changes in daily function, and six of the 15 papers reported a functional decline in a variable proportion of the population studied. Increased cognitive difficulties are directly related to more caregiver burden (Suárez-González et al., 2021). Adhering to IPC procedures also added to the increased burden of care for staff working with people living with dementia. The lived experiences of care home staff working with dementia patients highlighted the challenges of providing care when wearing PPE. For example, masks were sometimes ripped off the faces of care home staff by residents, which increased the risk of virus transmission despite staff adhering to IPC policies (Bunn, Brainard, Lane, Salter, & Lake, 2021). In the study by Bunn et al., (2020), care home staff also reported that IPC guidance was often unclear and frequently changing, which resulted in confusion and difficulty in adhering to guidelines.

A mixed method study by Tulloch et al., (2021) conducted in England, UK, investigated adherence to IPC and lateral flow testing. Rapid lateral flow tests were performed, but protocol adherence was poor, with 8.6% of staff achieving protocol adherence of >75% and 25.3% achieving ≥50%. The qualitative analysis revealed difficulties in implementing biweekly staff testing within already over-burdened care homes. Factors influencing adherence included excessive work burden, procedural and socio-economic factors, cognitive overload and the emotional impact of testing. To achieve protocol adherence, staff would need to sacrifice essential care duties. The opportunity costs of testing regimes need to be taken into consideration in future policymaking (Tulloch et al., 2021).

##### Increased burden of managing care

A cross-sectional survey by Sama et al., (2021) indicated the increased burden on domiciliary care managers due to COVID-19 IPC procedures. The majority of survey respondents, n=99 (85.1%) reported an increase in managers’ time spent developing new policies, procedures, and training (CI 95% 77.9-92.32). N=78 (67.0%) of the survey respondents reported an increase in the time managers spent scheduling (CI 95% 57.5-76.5) (Sama et al., 2021).

##### Absenteeism

Following a survey conducted in the USA by Rowe et al., (2020), it was reported that many caregivers (i.e., direct care workers, personal care assistants, home healthcare aides, personal attendants) “called off going into work” at the start of the COVID-19 pandemic. Reasons for absenteeism included caregiver illness, fears about COVID-19, competing responsibilities (e.g., childcare responsibilities), and confusion about federal and state stay-at-home orders. Home care agencies reported that many employees were quarantined due to COVID-19 illness and exposures. For example, one agency reported employees ‘calling off’ work because they have been exposed to the virus while at work in another job. Caregivers also feared COVID-19 exposure by caring for older adults, and many were categorised as high risk themselves (e.g., staff over the age of 65 with multiple comorbidities). Rowe et al., (2020) also noted that many of the initial new staff recruiting activities were postponed due to social distance precautions. Care home managers were also unable to do required background checks for new staff (Rowe et al., 2020).

**Bottom line. Increased IPC procedures during the COVID-19 pandemic increased stress and burden among care staff because of increased workload and dilemmas between adhering well to IPC procedures and providing the best care for the residents**.

#### 2.2.3 COVID-19 guidance and context

##### Policies, training and evaluation

The quantitative survey study by Rowe et al., (2020) was conducted in March 2020, at the beginning of the COVID-19 pandemic in the USA. They found that in March 2020, caregivers providing essential care for millions of older adults were largely absent from federal, state, and health system strategies for mitigating the spread of COVID-19. Rowe et al., (2020) noted that future policies must include health care agencies and to optimise care for older adults (Rowe et al., 2020).

In a mixed methods study conducted in England, Bunn et al., (2021) reported that care home staff adherence to IPC was high, but that fatalistic attitudes towards COVID-19 infection were also present (Bunn et al., 2021). Challenges of providing care using PPE, especially for residents with dementia, were highlighted. Interviewees reported dilemmas between the strict implementation of IPC procedures and conflicts with providing the best care to residents while preserving personal space. Nine months into COVID-19, official guidance was reported as confusing, constantly changing and poorly suited to care homes. Care home staff appreciated opportunities to work with other care homes and experts to interpret and implement guidance. IPA training was undertaken using multiple techniques but with little evaluation of these or how to sustain behaviour change.

##### A need for clearer pandemic plans

The qualitative study by Kaelen et al., (2021) also revealed the insights of care home residents and nursing home staff at the height of the early COVID-19 pandemic. Kaelen et al., (2021) suggested that clearer outbreak plans, including psychosocial support, could have prevented the aggravated mental health conditions of both residents and staff (Kaelen et al., 2021). They noted that a holistic approach is needed in nursing homes in which tailor-made essential restrictive IPC measures are combined with psychosocial support measures to reduce the impact on residents’ mental health and to enhance their quality of life. Similarly, Suarez-Gonzalez et al., (2021) found that lockdowns and confinement measures brought about by the COVID-19 pandemic have damaged the cognitive and psychological health and functional abilities of people living with dementia across the world. They recommend that safe care home visits can and should be maintained (Suárez-González et al., 2021).

##### Visitation

A mixed methods pilot study was conducted by Verbeek et al., (2021) in the Netherlands, which aimed to evaluate the impact of visitors being allowed back in nursing homes following the start of the COVID-19 pandemic (Verbeek et al., 2020). Descriptive results showed that, in total, during the first week of the pilot, 954 residents had received a visitor (57% of the total surveyed). Differences were observed in how nursing homes selected visitors. In total, 21 locations allowed visitors, in principle, for all of their residents. However, for 6 locations, visits were permitted for 80% or more of the residents. The other 15 locations only partially allowed visitors. Verbeek et al., (2021) found that compliance with local guidelines was sufficient to good (Verbeek et al., 2020). No new COVID-19 infections were reported during the time the study was conducted.

##### Rapid testing

A mixed methods study by Tulloch et al., (2021) in England, UK found that rapid testing had the potential to enable communication, to reopen care homes to visitors, and to gradually lift restrictions. In addition to the impact on family visits, restrictions have also limited visits from GPs and healthcare professionals (Tulloch et al., 2021). Tulloch et al., (2021) noted that the restoration of these visits, with increased healthcare support for residents, and healthcare advice for care home staff, would be a positive outcome (Tulloch et al., 2021).

**Bottom line. COVID-19 IPC procedures were not well developed at the beginning of the COVID-19 pandemic, but evidence from 2021 suggests that good adherence to IPC measures can enable visitations by family members and medical professionals into care homes**.

### 2.3 Adverse outcomes and harm reduction in domiciliary care

#### 2.3.1 Domiciliary care

Only one cross-sectional survey study by Sama et al., (2021) specifically reported domiciliary care outcomes relating to IPC procedures. This cross-sectional survey indicated the increased burden on domiciliary care managers due to COVID-19. The majority of survey respondents (85.1%) reported an increase in managers’ time spent developing new policies, procedures and training (CI 95% 77.9-92.32), and 67.0% reported an increase in the time managers spent scheduling (CI 95% 57.5-76.5) (Sama et al., 2021).

**Bottom line. Only one study investigating domiciliary care was found. Therefore, it is difficult to make conclusions related specifically to this care setting. This lack of evidence may partly be due to the restrictions on research staff from conducting face to face research with vulnerable groups during the COVID-19 pandemic**.

### 2.4 Cost-effectiveness and cost/budget impact considerations

None of the included papers investigated the costs or cost-effectiveness of the IPC measures in relation to the outcomes under consideration in this rapid review.

**Bottom line. No published studies have reported on the costs or cost-effectiveness of IPC measures or have explored the cost implications of adverse outcomes associated with IPC measures**.

## 3. DISCUSSION

### 3.1 Summary of the findings

A total of 15 papers were included in this rapid review; 14 were primary studies, including quantitative studies, qualitative studies and mixed methods studies, and one was a rapid review paper. The studies included research relating to adverse outcomes and harm reduction relating to staff and residents associated with IPC procedures and policies.

#### Adverse outcomes of COVID-19 IPC procedures on care residents

The studies identified in this rapid review have indicated that the mental health of care home residents has been negatively affected by the COVID-19 pandemic restrictions (El Haj et al., 2020; Van der Roest et al., 2020). An increase in depression and loneliness was reported among care home residents in two of the included studies (El Haj et al., 2020; Sizoo et al., 2020). Moreover, diminished wellbeing due to a loss of usual freedoms was also highlighted in the review by Suarez-Gonzales and colleagues (Suárez-González et al., 2021).

Clearer outbreak management plans, including psychosocial support, could have prevented the aggravated mental health conditions of both residents and staff (Bunn et al., 2021; Kaelen et al., 2021; Rowe et al., 2020; Telford et al., 2021). A holistic approach is needed in care homes in which tailor-made essential restrictive IPC measures are combined with psychosocial support measures to reduce the impact on residents’ mental health and to enhance their quality of life (Kaelen et al., 2021). The correct implementation of COVID-19 IPC policies and procedures could enable visitation in nursing homes (Telford et al., 2021).

#### Adverse outcomes of COVID-19 IPC procedures on care staff

Evidence was found in this rapid review that there was depression and anxiety amongst staff who felt unprepared for COVID-19 related challenges which included a lack of IPC guidelines, issues around PPE and unclear visitation policies (El Haj et al., 2020; Kaelen et al., 2021). There was also evidence that staff felt confronted with professional and ethical dilemmas and described being ‘trapped’ between IPC procedures and the care of the residents (Kaelen et al., 2021). Care home staff reported dilemmas between strictly implementing IPC procedures and preserving personal space in terms of social distancing from the residents (Bunn et al., 2021)).

Bunn et al., (2021) highlighted the conflict between implementing IPC procedures and providing the best care. Official guidance was reported as confusing, constantly changing, and poorly suited to care homes. Rowe et al., (2020) noted that future policies must include home care agencies and their caregivers to optimize care for older adults.

Reasons for care staff absenteeism are not fully understood but may be due to fear of getting ill with the COVID-19 virus and fear for the health of other family members. Shortages of healthcare staff may be due to the requirement for staff to receive the COVID-19 vaccine. However, this is not yet mandated by the UK Government. In addition, care home staff shortages may be a direct result of Brexit (Dalingwater, 2021).

Increased IPC procedures during the COVID-19 pandemic were associated with increased stress and burden among care home staff due to increased workload. In situations where COVID-19 incidence may rise again, it is important to recognise that there needs to be a balance between adhering to IPC procedures and providing the best care for care home residents and domiciliary care recipients. Issues such as adequate support structures for both staff and care recipients should be considered in future COVID-19 IPC policies.

### 3.2 Limitations of the available evidence

One limitation is the lack of high-quality evidence from the included studies. Confidence in the strength of evidence about adverse outcomes of COVID-19 IPC procedures was rated as ‘low’ overall. Whilst the majority of studies were deemed to be of moderate quality according to the quality appraisal checklists used, due to the nature of methods used and the standard of reporting, overall, the evidence was weak, and therefore quality was rated as ‘low’ overall.

The key limitations of included studies are summarised in this report. However, due to the rapid nature of this review, quality appraisals were conducted (see Appendix 1), but no formal risk of bias was conducted.

### 3.3 Implications for policy and practice

Since March 2020, there have been many changes to government guidelines relating to procedures to keep the population safe from COVID-19 harm. Policies vary according to country, even within the UK. Important issues such as care home visitation policies have changed in such a way that care home staff have felt it difficult to keep up with the changes, which in itself increased the burden on those staff. The implications identified from this work are as follows:

- Clear and concise IPC pandemic policies should be in place in every care home and domiciliary care settings
- Increased attention to workforce planning is needed to ensure adequate staffing and to reduce individual burden
- Restrictions (e.g., visitation) for care residents needs to be balanced by additional psychological support
- Further research with robust methods in this area is urgently needed especially in the domiciliary care setting

### 3.4 Strengths and limitations of this Rapid Review

#### Strengths

Where previous rapid reviews focused on the effectiveness of IPC procedures for reducing COVID-19 transmission in care homes, this rapid review focused on adverse outcomes experienced by residents and staff from IPC procedures in care homes and in domiciliary care. The behavioural outcomes included adherence to and confidence in IPC procedures, the burden of increased and changing IPC procedures, wellbeing and quality of life of staff and residents during various waves of the COVID-19 pandemic.

#### Limitations

Due to the various study designs, a quality appraisal was conducted with a variety of checklist tools, making comparisons difficult. Although efforts were made to include domiciliary care studies, only one study was found in this rapid review which investigated behavioural outcomes relating to changing IPC procedures in residential and domiciliary care. No cost studies have been published to investigate the relationship between IPC procedure costs and outcomes within residential or domiciliary care during the COVID-19 pandemic. The timing and speed of the review meant that the review team could only find published literature. Full integration/ synthesis of review findings could not be achieved due to heterogeneity of studies; therefore, a narrative synthesis was used.

Key limitations of included studies and reviews are summarised. However, due to the rapid nature of this review, no formal risk of bias was conducted.

## 4. RAPID REVIEW METHODS

### 4.1 Eligibility criteria

Eligibility criteria for the rapid review are shown in Table 1. This review includes all IPC measures and protocols in care homes and domiciliary care settings. The scope is limited to OECD countries with no other geographic limits applied. All study designs are eligible for inclusion in the review.

**Table 1:**
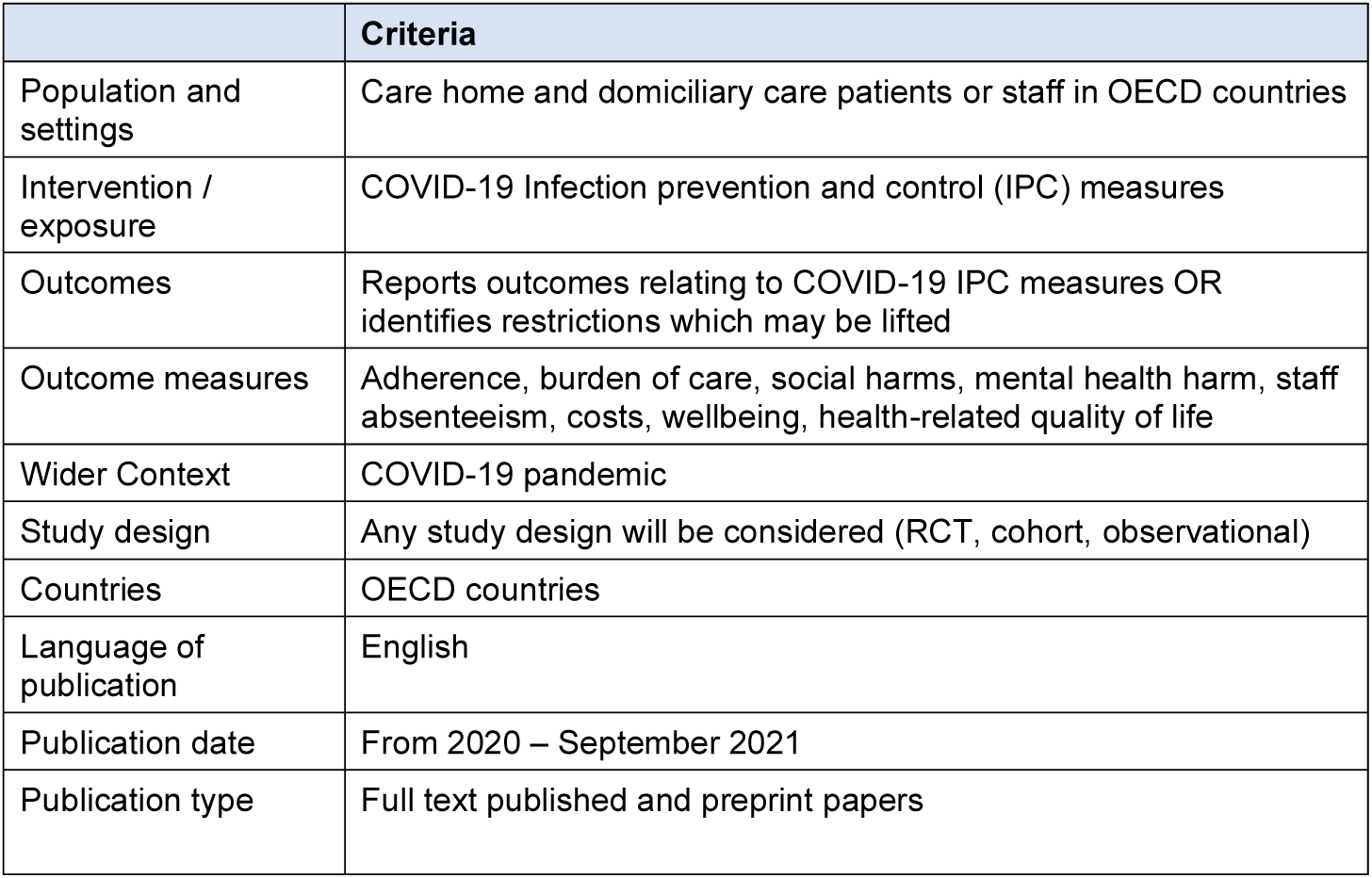
PICO for inclusion criteria.

**Table 2:**
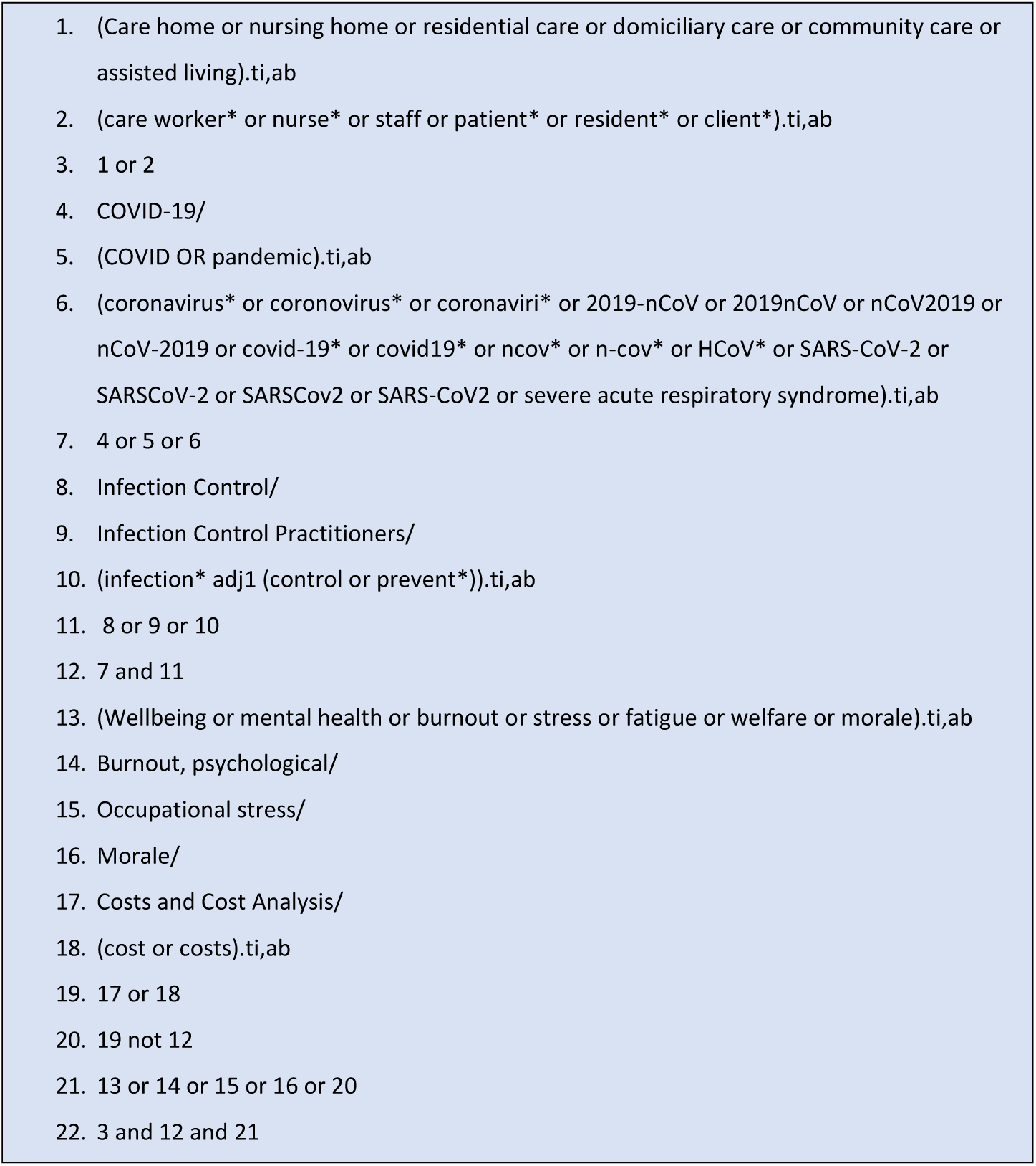
Search terms for MedLine (adapted for other databases)

### 4.2 Literature search

Key evidence sources include:

1. Medline
2. ASSIA
3. CINAHL
4. Cochrane Library
5. HTA libraries
6. Prospero

Key sources were searched for papers published between 1 January 2020 and 30 September 2021. The searches were limited to published research in the English language. The scope outlined for this search was to keep the review concise and deliverable within the time frame expected for a rapid review.

### 4.3 Study selection process

Key words included: wellbeing, costs, savings, buy-in, support, adherence, mental health, hospital admissions, care giver burden, deaths.

### 4.4 Reference management

The Covidence reference management system was used to store and manage citations. Duplicates were removed in Covidence (Veritas Health Innovation, 2021).

### 4.5 Data extraction

The data was extracted from the included studies using a pre-defined data extraction tool developed to capture all relevant data. Extracted data included study details such as author, year, setting, aim, design, population and sample size. The data extraction also includes data specific to the review question, type of study, method of analysis, key findings, and author conclusions.

Included papers were distributed among the review team for data extraction. A sample of extracted studies was checked against the papers for accuracy by the review lead. A proportion of the papers were double extracted to check for any discrepancies between reviewers.

A standardised data extraction table included the following categories:

- study citation (author, year of publication)
- study details (study aims, study design, geographical region, data collection dates)
- study methods (type of participants, sample size, outcome measures)
- study results
- additional notes

### 4.6 Assessment of body of evidence

Quality appraisal was carried out by members of the review team using the Mixed methods appraisal tool (MMAT) (Hong et al., 2018), the CEBM Survey quality appraisal checklist (Center for Evidence Based Management, 2005) and the following JBI critical appraisal tools: JBI analytical cross-sectional study checklist, JBI systematic reviews and research syntheses checklist, JBI case reports checklist, and JBI cohort studies checklist (Moola et al., 2020).

## 5. EVIDENCE

### 5.1 Study selection flow chart

The PRISMA flow chart (Page et al., 2021) is shown below:

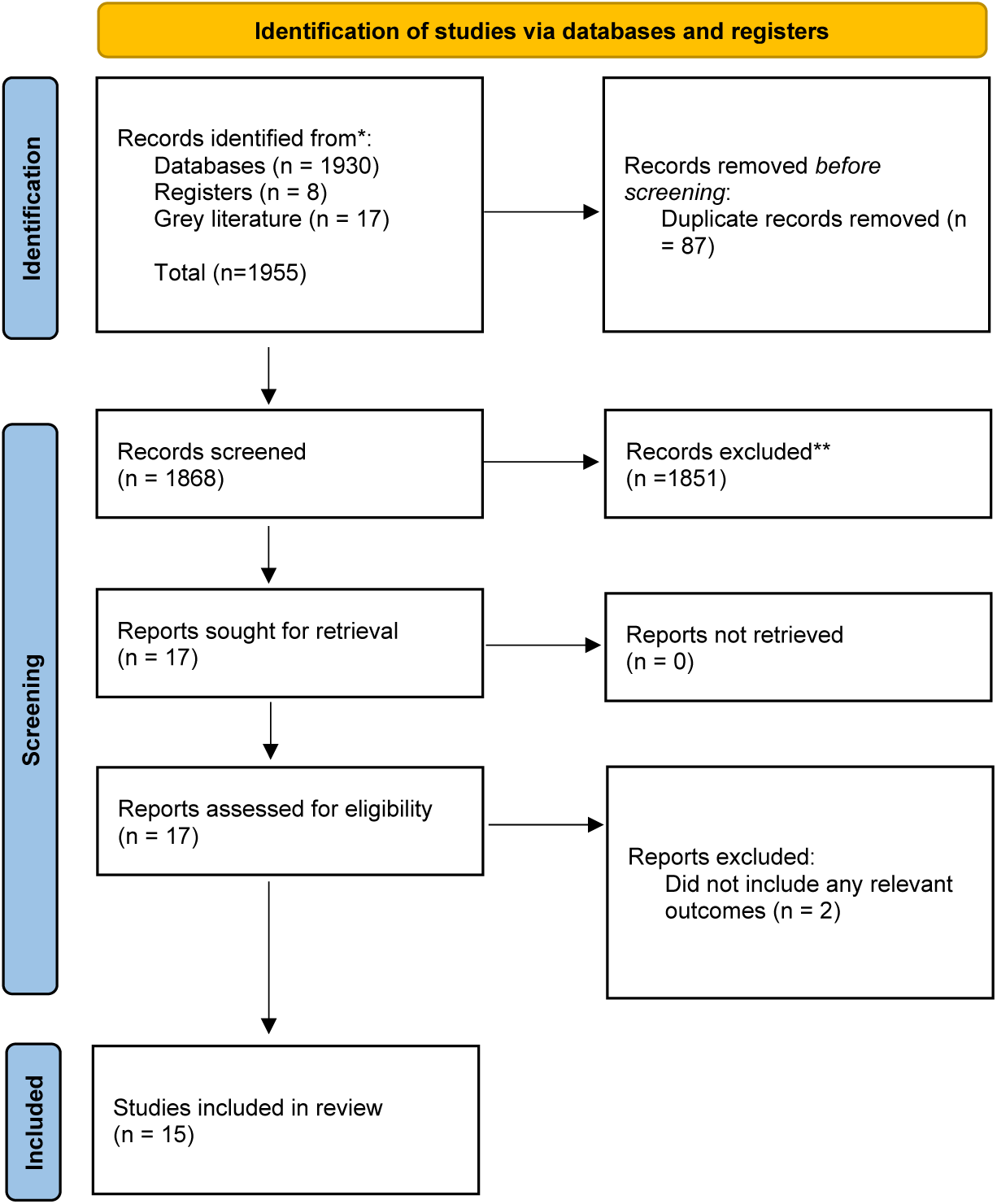
For more information, visit: http://www.prisma-statement.org/

### 5.2 Data extraction tables

The primary studies identified for this rapid review are detailed in Table 1.

**Table 1.** Primary studies included.

| Citation (Country) | Study Details | Participants & setting | Key findings | Observations/notes |
| --- | --- | --- | --- | --- |
| **Bunn et al., (2021) London, UK** | **Study Design:** Mixed methods<br>**Type of intervention:** Online survey and qualitative interviews to investigate the impact on care and staff morale.<br>**Data collection methods:** 238 online survey responses and 15 in-depth interviews with care home staff.<br>**Quality rating:** Moderate | **Participants:** 238 online survey responses and 15 in-depth interviews with care home staff.<br>**Setting:** Care homes in London, UK.<br>**Dates of data collection:** November 2020 to January 2021 in London, UK. | **Primary Findings:** Reported adherence to IPC was high, but fatalistic attitudes towards COVID-19 infection were present. Challenges of providing care using PPE, especially for residents with dementia, were highlighted. Care home staff reported dilemmas between strictly implementing IPC procedures and providing the best care to residents and preserving personal space.<br>Nine months into COVID-19, official guidance was reported as confusing, constantly changing and poorly suited to care homes. Care home staff appreciated opportunities to work with other care homes and experts to interpret and implement guidance.<br>IPA training was undertaken using multiple techniques but with little evaluation of these or how to sustain behaviour change. | This paper highlighted the conflict between implementing IPC procedures and providing the best care. Official guidance was reported as confusing, constantly changing, and poorly suited to care homes. The authors noted that clear, concise and care home focused IPC guidance was needed. |
| **El Haj et al., (2020)** | **Study Design:** Cohort study<br>The study aimed to assess depression and anxiety in participants with mild AD who live in retirement homes.<br>**Data collection methods:** On-site caregivers invited participants to complete the Hospital Anxiety and Depression Scale (Zigmond and Snaith, 1983).<br>**Quality rating:** Moderate | **Participants:** Fifty-eight participants with Alzheimer's disease living in retirement homes.<br>**Setting:** Retirement homes in France<br>**Dates of data collection:** Between March and May 2020 (after the start of the COVID-19 pandemic in March 2020 and before submission for publication in May 2020). | **Primary Findings:** Participants reported higher depression during ($M = 14.21$, $SD = 3.17$) than before ($M = 12.34$, $SD = 4.10$) the Covid-19 crisis ($Z = -2.84$, $p = .005$, Cohen's $d = 0.80$). Participants also reported higher anxiety during ($M = 13.24$, $SD = 3.39$) than before ($M = 11.38$, $SD = 4.36$) the Covid-19 crisis ($Z = -2.86$, $p = .004$, Cohen's $d = 0.81$). | The authors attributed the increase of depression and anxiety to the isolation of the residents and/or to the drastic changes in their daily life and the care they received. Decisions to reduce visitation and activities come at a cost to residents with Alzheimer's disease and their mental health. |
| **laboni et al., (2020) Canada** | **Study Design:** Case report<br>Case study of a person with dementia who is placed in COVID-19 quarantine on a dementia unit<br>**Data collection methods:** case study example<br>**Quality rating:** Moderate | **Participants:** A 65-year-old woman with frontotemporal dementia<br>**Setting:** Behavioural unit in a Nursing home in Canada<br>**Dates of data collection:** Between March 2020 and date of publication (August 2020). | **Primary Findings:**<br>The patient with frontotemporal dementia was isolated from the other care home residents due to testing positive for COVID-19. Her behaviour and strength necessitated the use of a pharmacological intervention. Her antipsychotic medication was changed to a more sedating agent, loxapine 5 mg BID, and increased to 10 mg BID 3 days later, with loxapine 5 mg used as needed. | Placing a patient with COVID-19 and dementia into isolation for 14 days did not result in any obvious physical or psychological consequences for the resident of a dementia unit in a nursing care home in Canada. |
| **Kaelen et al., (2021) Belgium** | **Study Design:** A qualitative study.<br>The study aimed to better understand psychosocial and mental health needs of nursing home residents during Covid-19 and how staff could respond accordingly.<br>**Data collection methods:** A qualitative study including focus groups and interviews. Thematic content analysis applied.<br>**Quality rating:** Moderate | **Participants:** Eight focus group discussions with direct care staff and 56 in-depth interviews with residents.<br>**Setting:** Nursing homes in Brussels, Belgium.<br>**Dates of data collection:** June 2020. | **Primary Findings:**<br>Nursing home residents experienced losses of freedom, social life, autonomy, and recreational activities that deprived them of their basic psychological needs, impacting mental wellbeing expressed in feeling depressed, anxious, and frustrated as well as decreased meaning and quality of life.<br>Staff felt unprepared for the challenges posed by the pandemic such as lacking guidelines, personal protective equipment and clarity around the organization of care. They were confronted with professional and ethical dilemmas, feeling 'trapped' between IPC and the residents' wellbeing. They witnessed the detrimental effects of the measures imposed on their residents. | Clearer outbreak management plans, including psychosocial support, could have prevented the aggravated mental health conditions of residents and staff. A holistic approach is needed in nursing homes in which tailor-made essential restrictive IPC measures are combined with psychosocial support to reduce the impact on residents' mental health and enhance their quality of life. |
| **Nestor et al. (2021), Ireland, UK** | **Study Design:** Cross-sectional survey<br>**Type of intervention:** Questionnaire to evaluate the impact of Covid-19 across a diverse range of care staff outcomes including wellbeing, workload and role definition.<br>**Data collection methods:** Survey<br>**Quality rating:** Moderate | **Sample size:** 250 respondents<br>**Participants:** Care staff including nurses and healthcare assistants<br>**Setting:** A combined elderly care and specialist palliative care facility. The elderly care unit provides care to long-term residents and short-term respite patients.<br>**Dates of data collection:** 6-week period commencing from the 11<sup>th</sup> September 2020 | **Primary Findings:**<br>182 (72.8%) respondents agreed that their responsibilities had increased during the pandemic. The majority of respondents (78.6%) reported an increase in work-related stress as a consequence of the Covid-19 pandemic. Constantly changing IPC protocols was ranked 4<sup>th</sup> of 17 highest rated workplace stressors by nursing and healthcare staff. Staff working exclusively in elderly care reported higher stress compared to the specialist palliative care workers in the following areas: | The study reported on outcomes across a combined palliative care and elderly care setting. It was not possible to attribute the findings solely to long-term elderly care staff. 38.9% of the survey respondents worked in the 'elderly care' setting. Of these, 21.2% worked across both elderly care and specialist palliative care settings. |
|  |  |  | • Challenge of adapting/ implementing changing workplace protocols (U = 1880, z = 2.74, p = 0.006)<br>• Suspending typical services normally available to patients (U = 1808, z = 3.29, p = 0.001)<br>• Wearing of masks/other PPE (U = 1991, z = 2.74, p = 0.006)<br>• Restricting patient access to pastoral support (U = 1726.50, z = 3.29, p = 0.001)<br>Failing to provide effective care because of time/staffing pressures (U = 1459.5, z = 3.63, p < 0.001). |  |
| **Ohta et al., (2021), Japan** | **Study Design:** Quantitative (cohort study)<br>Effect of COVID-19 on staff working conditions in rural nursing homes.<br>**Data collection methods:** Number of staff sickness absence days.<br>**Quality rating:** Moderate | **Participants:** Each group (pre- and post-COVID-19 control groups) had 48 participants.<br>**Setting:** Rural nursing homes in Japan<br>**Dates of data collection:** 1 April 2019 to 31 March 2021 | **Primary Findings:** The total number of days-off taken by the staff increased from the pre-COVID-19 to post-COVID-19 control period (79 vs. 155), but the difference was not statistically significant (p = 0.269). | Although more days were taken off post COVID-19 compared to pre-COVID-19, the difference was not statistically significant. |
| **Rowe et al., (2020), USA** | **Study Design:** Quantitative (survey)<br>The study aimed to characterize the effects of COVID-19 on Home Care Agencies (HCA) and examine HCAs' infection prevention and control (IPC) policies.<br>**Data collection methods:** Analysis of data collected from a national survey of HCAs. Surveys were e-mailed to members of a national HCA association on March 18, 2020. Questions included queries on demographics, COVID-19 exposures, effects of COVID-19, and IPC protocols | **Participants:** 1204 HCAs responded with an average census of 96.2 (5–2800) patients daily. Across 36 states, 238 HCAs reported COVID-19 cases or exposures among caregivers.<br>**Setting:** Home Care Agencies in the USA<br>**Dates of data collection:** The survey link was distributed on March 18, 2020, and closed on March 24, 2020. | **Primary Findings:** Due to COVID-19, many HCAs reported caregivers not coming to work or calling off. Reasons for absenteeism included caregiver illness, fears about COVID-19, competing responsibilities (e.g., childcare responsibilities), and confusion about federal and state stay-at-home orders. HCAs reported employees quarantined due to COVID-19 illness and exposures. Caregivers feared COVID-19 exposure by caring for older adults and many were high risk themselves (e.g. ages > 65 years, had multiple comorbidities).<br>**Additional Findings:** New staff recruiting activities were postponed. It was difficult to schedule orientations due to requisite hiring services | The authors reported that in March 2020, caregivers providing essential care for millions of older adults were largely absent from federal, state, and health system strategies for mitigating the spread of COVID-19. Future policies must include HCAs and caregivers to optimize care for older adults. |
|  | **Quality rating:** Moderate |  | being shut down. Care home managers were unable to do required background checks for new staff. |  |
| **Sama et al., (2021), USA** | **Study Design:** Cross-sectional survey to assess the impact of Covid-19 on home health and home care managers<br>**Data collection methods:** Online survey<br>**Quality rating:** Moderate | **Sample size:** 116 returned surveys (42% response rate)<br>**Participants:** Home Health and Home Care Agency Managers<br>**Dates of data collection:** Between 1<sup>st</sup> March and 30<sup>th</sup> June 2020 | **Primary Findings:** The majority of survey respondents (85.1%) reported an increase in managers' time spent developing new policies, procedures, and training (CI 95% 77.9-92.32).<br>67.0% reported an increase in the time managers spent scheduling (CI 95% 57.5-76.5). | This cross-sectional survey indicated the increased burden on domiciliary care managers due to COVID-19. |
| **Schweighart et al., (2021). Germany** | **Study Design:** Qualitative, semi-structured interviews<br>**Type of intervention [exposure]:** Nine interviews with nursing home residents. Depression screening was assessed using the 10 item "Depression in old age" scale<br>**Data collection methods:** Tape recorded interviews<br>**Quality rating:** Moderate | **Sample size:** 9 residents were interviewed<br>**Participants:** The survey included residents from inpatient nursing homes of whom a depressive illness was noted in the patient or who were judged to be depressed by the staff. Residents who, according to the patient file, had advanced dementia were excluded. At the time of the interviews, the residents were on average 81.9 years old.<br>**Setting:** Two Nursing Homes in Germany<br>**Dates of data collection:** December 2020 | **Primary Findings:** The average severity of depression was 5.9 points on the DIA-S. The value was slightly lower for residents with a documented diagnosis (M = 5.8) than for residents rated as depressed by the staff (M = 6.0). According to the DIA S, all of the respondents very likely- suffered from depression.<br>The residents reported increased boredom and inactivity, feeling cold due to ventilation requirements and isolation due to limited contact with other residents. They also found that wearing masks was a nuisance, uncomfortable to wear, created breathing difficulties, and caused glasses to fog. It was difficult to wear a hearing aid, glasses, and mask at the same time. | This study explored the experiences and perspectives of nursing home residents with depressive symptoms during the pandemic. Many adverse effects of IPC procedures were mentioned by the care home residents. |
| **Sizoo et al, (2020), The Netherlands** | **Study Design:** Qualitative, exploratory study to identify dilemmas experienced by Elderly Care Practitioners (ECPs) in daily practice as a result of the COVID-19 driven restrictive visiting policy in Dutch NHS<br>**Type of intervention:** The questions aimed to explore whether the dilemma related to the resident, | **Sample size:** The questionnaire was sent to 195 ECPs. 76 ECPs and ECPs-in-training completed the questionnaire describing 114 cases where they experienced a dilemma<br>**Participants:** ECPs and ECPs-in-training<br>**Setting:** Amsterdam University Medical Centre<br>**Dates of data collection:** Between April 17 and May 10, 2020 | **Primary Findings:** ECPs observed loneliness, depressive symptoms, decreased intake, increase in somatic symptoms (i.e., pain), physical deterioration in residents.<br>In psychogeriatric residents', ECPs observed rapid cognitive decline, and changes in neuropsychiatric symptoms, including agitation and aggression. | An open-ended questionnaire was outlined as a pragmatic study design to gather qualitative data that allowed respondents to reply at their own convenience. In addition, respondents might also be prone to more honest answers in an anonymous survey. |
|  | the resident's family, the nursing staff, care unit, and/or organization.<br>**Data collection methods:** An open-ended questionnaire based on discussions on the impact of the COVID-19 driven restrictive visiting policy.<br>**Quality rating:** Moderate |  | The latter was even reported to result in increased drug prescriptions for some of the residents.<br>ECPs felt moral issues with adhering to IPC measures. ECPs described emotions of guilt and injustice.<br>ECPs considered the presence of visitors as essential to quality of life. As most residents in nursing homes have limited life expectancy, ECPs estimated quality of life was often considered more important than life duration. |  |
| **Telford et al., (2021), USA** | **Study Design:** Quantitative (cross-sectional study)<br>**Data collection methods:** Site visits to 24 long term care facilities. Thirty-three key indicators were evaluated from five Infection Prevention Control (IPC) categories: Hand Hygiene, Disinfection, Social Distancing, Personal Protection Equipment (PPE) and Screening for symptoms and elevated temperature.<br>**Quality rating:** Moderate | **Sample size:** 24 long term care facilities were evaluated for key indicators of infection prevention control<br>**Participants:** 2,580 residents in 24 long term care facilities<br>**Setting:** Long term care facilities in the USA<br>**Dates of data collection:** June-July 2020 | **Primary Findings:** Long term care facilities with lower COVID-19 prevalence among residents had significantly greater implementation of IPC procedures compared to those with higher COVID-19 prevalence, suggesting the utility in adhering to current guidelines to reduce transmission.<br>**Additional Findings:** IPC implementation among higher-prevalence COVID-19 long term care facilities (LTCF) was significantly weaker than lower prevalence LTCFs in the "Social Distancing" and PPE categories. | Better adherence to IPC procedures led to better outcomes in terms of fewer COVID-19 transmissions. |
| **Tulloch et al., (2021), UK** | **Study Design:** Mixed methods<br>**Type of intervention:** Evaluation of lateral flow device-based testing protocol.<br>**Data collection methods:** Descriptive epidemiological analysis of COVID-19 testing, case data and qualitative interviews<br>**Quality rating:** High | **Sample size:** 11 care homes included in the quantitative analysis. Qualitative exploration was conducted in 9 of the care homes<br>**Participants:** Care home staff<br>**Setting:** Care homes in Liverpool, England, UK<br>**Dates of data collection:** Between 1st December 2020 and 10th January 2021 | In total, 1,638 LFD rapid tests were performed on 407 staff. Protocol adherence was poor with 8.6% of staff achieving protocol adherence of >75%, and 25.3% achieving ≥50%.<br>Qualitative analysis revealed difficulties in implementing testing strategies due to excessive work burden. Factors influencing adherence related to test integration and procedural factors, socio-economic factors, cognitive overload and the emotional value of testing. | Adherence to testing protocol was poor among care home staff. |
| **Van der Roest et al. (2020), The Netherlands** | **Study Design:**<br>Cross-sectional survey<br>**Type of intervention:**<br>The impact of Covid-19 measures on wellbeing of older long-term care facility (LTCF) residents<br>**Data collection methods:**<br>Semi-open online survey<br>**Quality rating:** Low | **Sample size:**<br>193 long term care facility residents<br>**Participants:**<br>3 independent samples of residents without severe cognitive impairment (CI), family members of residents with and without CI, and care staff from all unit-types in Dutch LTCFs (nursing homes and residential care facilities)<br>**Dates of data collection:**<br>Between 30<sup>th</sup> April and 27<sup>th</sup> May 2020 | Mood in residents was assessed with the Mental Health Inventory 5-index (MHI-5; range 0e100, scores <60 indicate poor mental health). Mean MHI-5 score for residents was 56.6 (SD 20.4), 51% had scores <60. Happiness was less often, and sadness was more often reported by families of residents without CI than with CI (P < .000; P < .008, respectively). Loneliness was reported by 149 (77%) residents. Staff classified residents without CI lonelier than residents with CI (P < .006). More than half of the staff reported an increase in severity of agitation, depression, anxiety, and irritability. | The authors concluded that during the COVID-19 measures, wellbeing of older LTCF residents was severely affected. Due to the visitor ban, high levels of loneliness, depression, and a significant exacerbation in mood and behavioural problems were reported. Residents without CI seemed to be the most affected. |
| **Verbeek et al., (2020), The Netherlands** | **Study Design:** A mixed methods cross-sectional study evaluating the impact of visitors being allowed back in nursing homes following the start of the COVID-19 pandemic.<br>**Data collection methods:**<br>Questionnaire, telephone interviews, analyses of documentation ( i.e.,local visiting protocols), and a WhatsApp group.<br>**Quality rating:** Moderate | **Participants:** Telephone interviews were conducted with contact persons of 26 nursing homes (100%), 24 electronic questionnaires were returned (92%), and for 23 nursing homes (88%) documentation, including local protocols, was received. In total, 30 persons participated in the WhatsApp group, representing 20 nursing homes (77%), of which 4 nursing homes were represented by 2 persons, and 3 nursing homes were represented by 3 persons.<br>**Setting:** Nursing homes in The Netherlands<br>**Dates of data collection:** Between 11<sup>th</sup> May and 5<sup>th</sup> June 2020. | **Primary Findings:**<br>Descriptive results showed that during the first week of the study, 954 residents had received a visitor (57%). Differences were observed in how nursing homes selected visitors. In total, 21 locations allowed visitors for all of their residents. However, for 6 locations visits were permitted for 80% or more of the residents. The other 15 locations only partially allowed visitors, varying from 20% to 70% of residents.<br>**Additional Findings:**<br>Compliance with local guidelines was sufficient to good.<br>**Additional Findings:** No new COVID-19 infections were reported during the time the study was conducted. | All nursing homes recognized the added value of real and personal contact between residents and their visiting loved ones and indicated a positive impact on wellbeing. Based on these results, the Dutch government decided to allow all nursing homes in the Netherlands to cautiously open their homes using the 2020 IPC guidelines. |

**Table 2:** Summary of included review paper.

| Citation (Country) | Review details | Included studies | Quality | Findings and observations/notes |
| --- | --- | --- | --- | --- |
| Suarez-Gonzalez et al., (2021), UK and India | **Type of review:** Rapid review (with one year of evidence)<br>**Review period:** Up to February 2021.<br>**Review purpose:** The review aimed to examine and summarise the global research evidence describing the effect of COVID-19 isolation measures on the health of people living with dementia worldwide.<br>**Included study designs:** Qualitative and quantitative<br>**Quality rating:** Moderate | **Number of included studies:** 15 papers included<br>**Key characteristics:**<br>People with dementia living in care homes and within the community. Changes were observed for the following categories:<br>Cognition<br>Behavioural symptoms<br>Psychological symptoms<br>Daily function | **Primary Findings:** 15 eligible papers, examining a total of 6,442 people with dementia. 13/15 were conducted in people living in the community and 2 in care homes. 60% (9/15) of studies reported changes in cognition with 77% (7/9) describing declined cognition by >50% of respondents. 93% (14/15) of studies reported worsening or new onset of behavioural and psychological symptoms. 46% (7/15) studies reported changes in daily function, and 6 papers reported a functional decline in a variable proportion of the population studied. | Lockdowns and confinement measures brought about by the COVID-19 pandemic have damaged the cognitive and psychological health and functional abilities of people with dementia across the world. Safe care home visits should be maintained along with adherence to IPC procedures. Increased cognitive difficulties relates to more caregiver burden. Adhering to IPC procedures adds to the increased burden of care. |

## Data Availability

All data produced in the present study are available upon reasonable request to the authors

## Abbreviations

Acronym/Abbreviation: Full description
BAME: Black, Asian and Minority Ethnic
Brexit: Brexit is the name given to the United Kingdom’s departure from the European Union. It is a combination of Britain and exit
GP: General Practitioner
HCA: Health Care Agencies
HCEC: Health and Care Economics Cymru
IPAC: Infection Prevention and Control
IPC: Infection Prevention Control
JBI: Joanna Briggs Institute
LFD: Lateral Flow Devices
LTCF: Long Term Care Facility
M: Mean
NHS: National Health Service
NICE: National Institute for Health and Care Excellence
OECD: Organisation for Economic Co-operation and Development
PCR: test Polymerase Chain Reaction test
PHE: Public Health England
PHW: Public Health Wales
PICO framework: Participant, Intervention, Comparison, Outcomes framework
PPE: Personal Protective Equipment
RCT: Randomised Controlled Trial
RES: Rapid Evidence Summary
SCIE: Social Care Institute for Excellence
SD: Standard Deviation
TAG: Technical Advisory Group
WC19EC: Wales COVID-19 Evidence Centre

## 7.2 Acknowledgements and author contributions

The authors would like to thank Marion Lyons and Angela Street for acting as expert stakeholders; Dr Ruth Lewis and Dr Alison Cooper for providing support to the review team; and Nicola Pearce-Smith, Public Health England for reviewing our search strategy.

## Author contributions

Project leads: CW, RTE, NB, DH; drafting of report: LHS, RTE, NH; contribution to writing and critical editing of the report; LHS, NH, AH, BA, AM, NN, NB CW, RTE, DH, DF; reviewing: LHS, BA, NH, AH, AM.

## 8. ABOUT THE WALES COVID-19 EVIDENCE CENTRE (WCEC)

The WCEC integrates with worldwide efforts to synthesise and mobilise knowledge from research.

We operate with a core team as part of Health and Care Research Wales, are hosted in the [Wales Centre for Primary and Emergency Care Research (PRIME)](http://www.primecentre.wales/), and are led by [Professor Adrian Edwards of Cardiff University](https://www.cardiff.ac.uk/people/view/123022-edwards-adrian).

The core team of the centre works closely with collaborating partners in [Health Technology Wales](https://www.healthtechnology.wales/), Wales Centre for Evidence-Based Care, [Specialist Unit for Review Evidence centre](https://www.cardiff.ac.uk/research/explore/research-units/wales-centre-for-evidence-based-care), [SAIL Databank](https://healthandcareresearchwales.org/about-research-community/secure-anonymised-information-linkage-sail-databank), [Bangor Institute for Health & Medical Research](https://www.bangor.ac.uk/health-sciences/research/index.php.en)/ Health and Care Economics Cymru, and the [Public Health Wales Observatory](https://phw.nhs.wales/services-and-teams/observatory/).

Together we aim to provide around 50 reviews per year, answering the priority questions for policy and practice in Wales as we meet the demands of the pandemic and its impacts.

Director:

Professor Adrian Edwards

Contact

**Website: https://healthandcareresearchwales.org/about-research-community/wales-covid-19-evidence-centre**

### 9. APPENDIX

#### Appendix 1 Quality appraisal tables

Members of the review team chose the most appropriate JBI critical appraisal tool. A quarter of critical appraisals will be checked by a second reviewer. Discrepancies arising during the critical appraisal process will be discussed until an agreement is reached by the review team. When possible, studies will be graded as ‘very low’, ‘low’, ‘moderate’ or ‘high’ quality.

**Table 4.** *JBI analytical cross-sectional study checklist* (Joanna Briggs Institute, 2017a)

| Citation | Q1. Were the criteria for inclusion in the sample clearly defined? | Q2. Were the study subjects and the setting described in detail? | Q3. Was the exposure measured in a valid and reliable way? | Q4. Were objective, standard criteria used for measurement of the condition? | Q5. Were confounding factors identified? | Q6. Were strategies to deal with confounding factors stated? | Q7. Were the outcomes measured in a valid and reliable way? | Q8. Was appropriate statistical analysis used? |
| --- | --- | --- | --- | --- | --- | --- | --- | --- |
| Telford et al., (2021) | Yes | Yes | Yes | Yes | Yes | No | Yes | Yes |

**Table 5.** *JBI Systematic Reviews and Research Syntheses Checklist* (Joanna Briggs Institute, 2017b)

| Citation | Q1. Is the review question clearly and explicitly stated? | Q2. Were the inclusion criteria appropriate for the review question? | Q3. Was the search strategy appropriate? | Q4. Were the sources and resources used to search for studies adequate? | Q5. Were the criteria for appraising studies appropriate? | Q6. Was critical appraisal conducted by two or more reviewers independently? | Q7. Were there methods to minimize errors in data extraction? | Q8. Were the methods used to combine studies appropriate? | Q9. Was the likelihood of publication bias assessed? | Q10. Were recommendations for policy and/or practice supported by the reported data? | Q11. Were the specific directives for new research appropriate? |
| --- | --- | --- | --- | --- | --- | --- | --- | --- | --- | --- | --- |
| Suarez-Gonzalez et al., (2021) | Yes | Yes | Unclear | Unclear | Yes | Yes | Yes | Yes | No | Yes | Unclear |

**Table 6.**
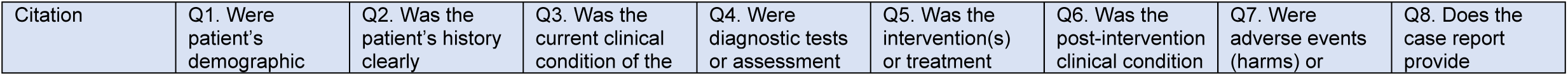
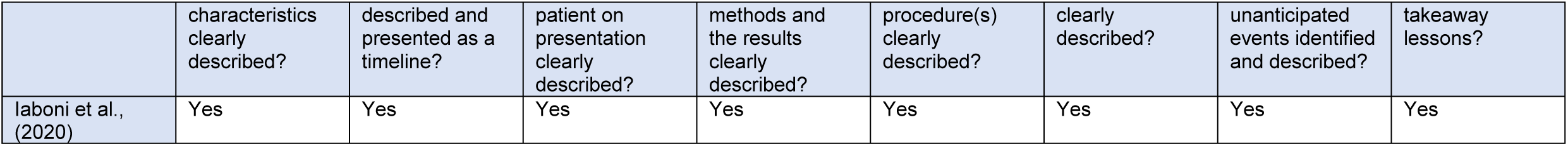
JBI Critical Appraisal Checklist for Case Reports

**Table 7.** JBI Critical Appraisal Checklist for Cohort Studies

| Citation | Q1. Were the two groups similar and recruited from the same population? | Q2. Were the exposures measured similarly to assign people to both exposed and unexposed groups? | Q3. Was the exposure measured in a valid and reliable way? | Q4. Were confounding factors identified? | Q5. Were strategies to deal with confounding factors stated? | Q6. Were the groups/ participants free of the outcome at the start of the study (or at the moment of exposure)? | Q7. Were the outcomes measured in a valid and reliable way? | Q8. Was the follow up time reported and sufficient to be long enough for outcomes to occur? | Q9. Was follow up complete, and if not, were the reasons to loss to follow up described and explored? | Q10. Were strategies to address incomplete follow up utilized? | Q11. Was appropriate statistical analysis used? |
| --- | --- | --- | --- | --- | --- | --- | --- | --- | --- | --- | --- |
| Ohta et al., (2021) | Yes | Yes | Yes | Yes | Yes | N/A | Yes | Yes | N/A | N/A | Yes |

**Table 8.** *JBI analytical quasi-experimental checklist* (Joanna Briggs Institute, 2021)

| Citation | Q1 Is it clear in the study what is the 'cause' and what is the 'effect' (i.e. there is no confusion about which variable comes first)? | Q2 Were the participants included in any comparisons similar? | Q3 Were the participants included in any comparisons receiving similar treatment/care, other than the exposure or intervention of interest? | Q4 Was there a control group? | Q5 Were there multiple measurements of the outcome both pre and post the intervention/exposure? | Q6 Was follow up complete and if not, were differences between groups in terms of their follow up adequately described and analysed? | Q7 Were the outcomes of participants included in any comparisons measured in the same way? | Q8 Were outcomes measured in a reliable way? | Q9 Was appropriate statistical analysis used? |
| --- | --- | --- | --- | --- | --- | --- | --- | --- | --- |
| El Haj et al., (2020) | Yes | No | No | No | No | Yes | Yes | Yes | Yes |

**Table 9.** *JBI analytical qualitative checklist* (The Joanna Briggs Institute, 2017)

| Citation | Q1 Is there congruity between the stated philosophical perspective and the research methodology? | Q2 Is there congruity between the research methodology and the research question or objectives? | Q3 Is there congruity between the research methodology and the methods used to collect data? | Q4 Is there congruity between the research methodology and the representation and analysis of data? | Q5 Is there congruity between the research methodology and the interpretation of results? | Q6 Is there a statement locating the researcher culturally or theoretically? | Q7 Is the influence of the researcher on the research, and vice-versa, addressed? | Q8 Are participants, and their voices, adequately represented? | Q9 Is the research ethical according to current criteria or, for recent studies, and is there evidence of ethical approval by an appropriate body? | Q10 Do the conclusions drawn in the research report flow from the analysis, or interpretation, of the data? |
| --- | --- | --- | --- | --- | --- | --- | --- | --- | --- | --- |
| Kaelen et al., (2021) | Yes | Yes | Yes | Yes | Yes | No | No | Yes | Yes | Yes |
| Schweighart et al., (2021) | Yes | Yes | Yes | Yes | Yes | No | No | Yes | Yes | Yes |
| Sizoo et al., (2020) | Yes | Yes | Yes | Yes | Yes | No | No | Yes | No | Yes |

**Table 10.** *CEBM Survey quality appraisal checklist* (Center for Evidence Based Management, 2005)

| Citation | Q1 Did the study address a clearly focused question / issue? | Q2 Is the research method (study design) appropriate for answering the research question? | Q3 Is the method of selection of the subjects (employees, teams, divisions, organizations) clearly described? | Q4 Could the way the sample was obtained introduce (selection) bias? | Q5 Was the sample of subject's representative with regard to the population to which the findings will be referred? | Q6 Was the sample size based on pre-study considerations of statistical power? | Q7 Was a satisfactory response rate achieved? | Q8 Are the measurements (questionnaires) likely to be valid and reliable? | Q9 Was the statistical significance assessed? | Q10 Are confidence intervals given for the main results? | Q11 Could there be confounding factors that haven't been accounted for? | Q12 12. Can the results be applied to your organization? |
| --- | --- | --- | --- | --- | --- | --- | --- | --- | --- | --- | --- | --- |
| Nestor et al., (2021) | Yes | Yes | Yes | No | Yes | No | Yes | Yes | Yes | No | No | Yes |
| Rowe et al., (2020) | Yes | Yes | Yes | No | Yes | N/A | Yes | Yes | Yes | No | No | N/A |
| Sama et al. 2021 | Yes | Yes | Yes | Yes | Yes | No | Yes | Unclear | No | Yes | No | Yes |
| Van der Roest et al. 2020 | Yes | Yes | No | Unclear | Unclear | No | Unclear | Yes | Yes | No | Unclear | Yes |

**Table 11.**
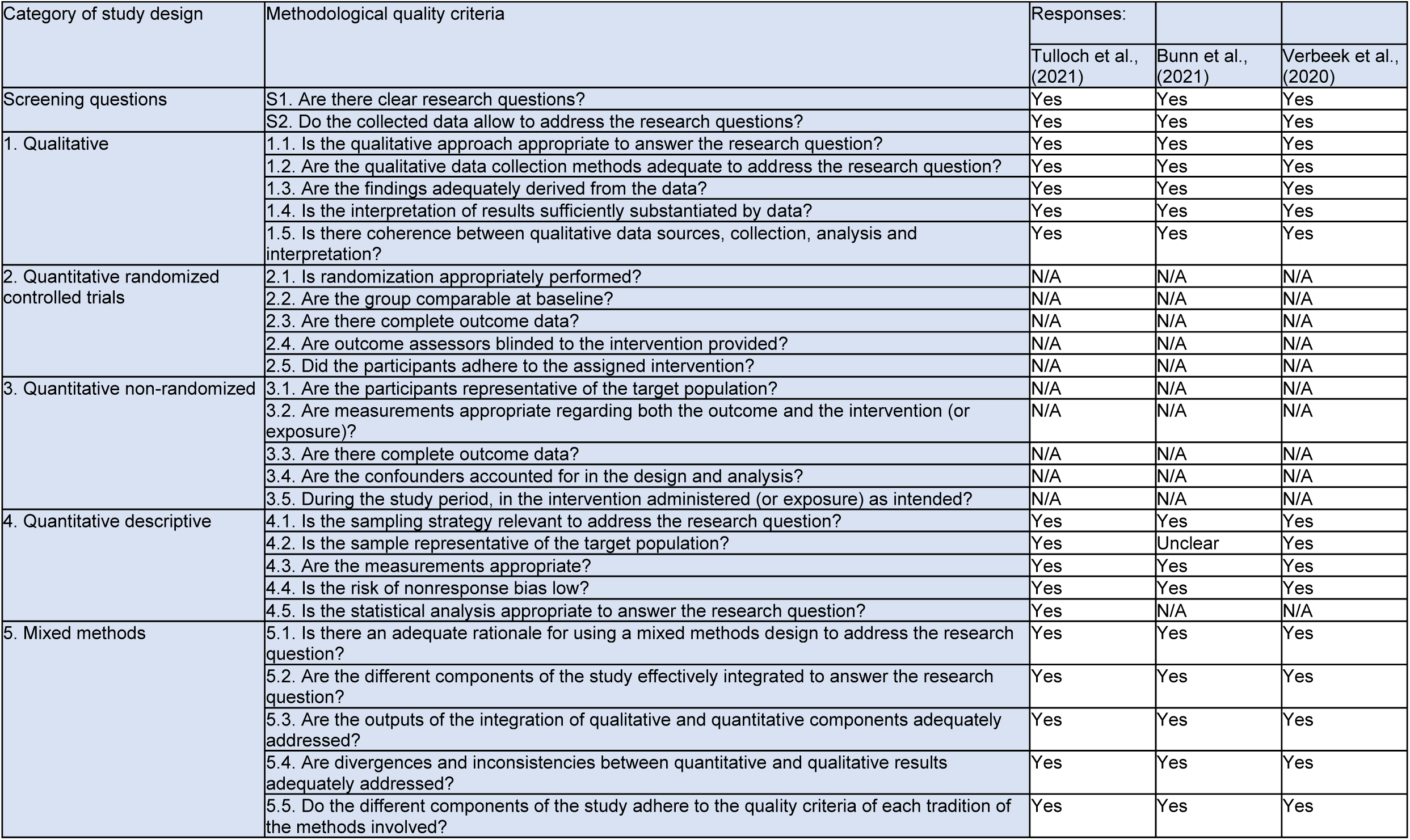
*Mixed methods appraisal tool (MMAT)* (Hong et al., 2018)

## REFERENCES

Bell, D., Comas-Herrera, A., Henderson, D., Jones, S., Lemmon, E., Moro, M., … Patrignani, P. (2020). COVID-19 mortality and long-term care: a UK comparison. Retrieved from https://ltccovid.org/wp-content/uploads/2020/08/COVID-19-mortality-in-long-term-care-final-Sat-29-1.pdf

Bunn, D., Brainard, J., Lane, K., Salter, C., & Lake, I. (2021). The Lived Experience of Implementing Infection Control Measures in Care Homes during two waves of the COVID-19 Pandemic. A mixed-methods study. MedRxiv.

Center for Evidence Based Management. (2005). Critical Appraisal of a Survey: Appraisal questions. Retrieved from http://www.cebma.org/wp-content/uploads/Critical-Appraisal-Questions-for-a-Survey.pdf

Dalingwater, L. L. (2021). NHS staffing shortages and the Brexit effect. Retrieved from http://journals.openedition.org/osb, (2019), 67-86, (24)

Department of Health & Social Care. (2021). Adult Social Care Extension to Infection Control and Testing Fund Ring-Fenced Grant 2021.

Duval, D., Sadler, L., Walters, B., Hooper, L., Pearce-Smith, N., & Clark, R. (2021). Interventions to reduce COVID-19 transmission in adult social care settings: a rapid review. UKHSA COVID-19 Rapid Evidence Service. 2021.

El Haj, M., Altintasb, E., Chapelete, G., Kapogiannisg, D., & Galloujb, K. (2020). High depression and anxiety in people with Alzheimer’s disease living in retirement homes during the covid-19 crisis. Psychiatry Research, 291(January), 19–21.

Groenewold, M. R., Burrer, S. L., Ahmed, F., Uzicanin, A., Free, H., & Luckhaupt, S. E. (2020). Increases in Health-Related Workplace Absenteeism Among Workers in Essential Critical Infrastructure Occupations During the COVID-19 Pandemic — United States, March–April 2020. MMWR. Morbidity and Mortality Weekly Report, 69(27), 853– 858. https://doi.org/10.15585/mmwr.mm6927a1

Hong, Q. N., Pluye, P., Fàbregues, S., Bartlett, G., Boardman, F., Cargo, M., … Vedel, I. (2018). Mixed Methods Appraisal Tool (MMAT), Version 2018. User guide. McGill, 1– 11. Retrieved from http://mixedmethodsappraisaltoolpublic.pbworks.com/w/file/fetch/127916259/MMAT_2018_criteria-manual_2018-08-01_ENG.pdf%0Ahttp://mixedmethodsappraisaltoolpublic.pbworks.com/

House of Commons. (2021). Workforce burnout and resilience in the NHS and social care -Health and Social Care Committee - House of Commons. Retrieved from https://publications.parliament.uk/pa/cm5802/cmselect/cmhealth/22/2206.htm

House of Commons Committees. (2021). How can we tackle staff burnout in the health and care sectors? Retrieved from https://houseofcommons.shorthandstories.com/health-and-care-staff-burnout/index.html

House of Commons, Health and Social Care, A., Technology, S. and, & Committees. (2021). Coronavirus: Lessons learned to date. Retrieved from https://committees.parliament.uk/publications/7496/documents/78687/default/

Iaboni, A., Cockburn, A., Marcil, M., Rodrigues, K., Marshall, C., Garcia, M. A., … Flint, A. J. (2020). Achieving Safe, Effective, and Compassionate Quarantine or Isolation of Older Adults With Dementia in Nursing Homes. American Journal of Geriatric Psychiatry, 28(8), 835–838. https://doi.org/10.1016/j.jagp.2020.04.025

Iacobucci, G. (2020). Covid-19: Lack of PPE in care homes is risking spread of virus, leaders warn. BMJ (Clinical Research Ed.), 368(March), m1280. https://doi.org/10.1136/bmj.m1280

Joanna Briggs Institute. (2017a). Checklist for Analytical Cross Sectional Studies. Joanna Briggs Institute Reviewer’s Manual, 6.

Joanna Briggs Institute. (2017b). Checklist for Systematic Reviews and Research Syntheses. The Joanna Briggs Institute. Retrieved from http://joannabriggs.org/research/critical-appraisal-tools.htmlwww.joannabriggs.org%0Awww.joannabriggs.org

Joanna Briggs Institute. (2021). Checklist for quasi-experimental studies (non-randomised experimental studies). Retrieved from https://jbi.global/critical-appraisal-tools

Kaelen, S., van den Boogaard, W., Pellecchia, U., Spiers, S., de Cramer, C., Demaegd, G., … Draguez, B. (2021). How to bring residents’ psychosocial wellbeing to the heart of the fight against Covid-19 in Belgian nursing homes - A qualitative study. PLoS ONE, 16(3 March), 1–19. https://doi.org/10.1371/journal.pone.0249098

Kavanagh, A., Hatton, C., Stancliffe, R. J., Aitken, Z., King, T., Hastings, R., … Emerson, E. (2021). Health and healthcare for people with disabilities in the UK during the COVID-19 pandemic. Disability and Health Journal, (xxxx), 101171. https://doi.org/10.1016/j.dhjo.2021.101171

Lauter, S., Lorenz-Dant, K., Perobelli, E., Caress, A., Sinha, S. K., Arling, G., & Comas-Herrera, A. (2021). International “living” report: LongTerm Care and COVID-19 vaccination, prioritization and data. Ltccovid.Org, (January), 1–11. Retrieved from https://ltccovid.org/wp-content/uploads/2021/01/COVID-19-vaccine-and-LTC-prioritization-and-data-26-January-3.pdf

Lebrasseur, A., Fortin-Bédard, N., Lettre, J., Bussières, E. L., Best, K., Boucher, N., … Routhier, F. (2021). Impact of COVID-19 on people with physical disabilities: A rapid review. Disability and Health Journal, 14(1). https://doi.org/10.1016/j.dhjo.2020.101014

Lipsitz, L. A., Lujan, A. M., Dufour, A., Abrahams, G., Magliozzi, H., Herndon, L., & Dar, M. (2020). Stemming the Tide of COVID-19 Infections in Massachusetts Nursing Homes. Journal of the American Geriatrics Society, 68(11), 2447–2453. https://doi.org/10.1111/jgs.16832

Low, L.-F., Hinsliff-Smith, K., Sinha, S., Stall, N., Verbeek, H., Siette, J., … Comas-Herrera, A. (2021). Safe visiting at care homes during COVID-19: A review of international guidelines and emerging practices during the COVID-19 pandemic, (January), 1–22. Retrieved from LTCcovid.org

Moola, S., Z, Munn Z.,, Tufanaru, Z., Aromataris, E., Sears, K., Sfetcu, R., … Mu, P.-F. (2020). JBI Manual for Evidence Synthesis. In A.E & Z. Munn (Eds.), 2020. Retrieved from https://synthesismanual.jbi.global

Morciano, M., Stokes, J., Kontopantelis, E., Hall, I., & Turner, A. J. (2021). Excess mortality for care home residents during the first 23 weeks of the COVID-19 pandemic in England: a national cohort study. BMC Medicine, 19(1), 1–11. https://doi.org/10.1186/s12916-021-01945-2

Nestor, S., O’ Tuathaigh, C., & O’ Brien, T. (2021). Assessing the impact of COVID-19 on healthcare staff at a combined elderly care and specialist palliative care facility: A cross-sectional study. Palliative Medicine, 35(8), 1492–1501. https://doi.org/10.1177/02692163211028065

NHS Confederation. (2021). A reckoning: the continuing cost of COVID-19 Overview. NHS Providers, (September), 1–24. Retrieved from https://ifs.org.uk/publications/15432

NHS National Services Scotland. (2021). Rapid Review of the literature: Assessing the infection prevention and control measures for the prevention and management of COVID-19 in health and care settings. Retrieved from https://hpspubsrepo.blob.core.windows.net/hps-website/nss/2985/documents/1_covid-19-rapid-review-ipc-for-covid-19.pdf

Niedzwiedz, C. L., O’Donnell, C. A., Jani, B. D., Demou, E., Ho, F. K., Celis-Morales, C., … Katikireddi, S. V. (2020). Ethnic and socioeconomic differences in SARS-CoV-2 infection: Prospective cohort study using UK Biobank. BMC Medicine, 18(1), 1–14. https://doi.org/10.1186/s12916-020-01640-8

Office for National Statistics. (2021a). Deaths involving COVID-19 in the care sector, England and Wales: deaths registered between week ending 20 March 2020 and week ending 2 April 2021. Retrieved from https://www.ons.gov.uk/peoplepopulationandcommunity/birthsdeathsandmarriages/deaths/articles/deathsinvolvingcovid19inthecaresectorenglandandwales/latest

Office for National Statistics. (2021b). Updated estimates of coronavirus (COVID-19) related deaths by disability status, England: 24 January to 20 November 2020.

Ouslander, J. G., & Grabowski, D. C. (2020). COVID-19 in Nursing Homes: Calming the Perfect Storm. Journal of the American Geriatrics Society, 68(10), 2153–2162. https://doi.org/10.1111/jgs.16784

Page, M., McKenzie, J., Bossuyt, P., Boutron, I., Hoffmann, T., & Mulrow, C. (2021). The PRISMA 2020 statement: an updated guideline for reporting systematic reviews. BMJ 2021. BMJ. https://doi.org/doi:10.1136/bmj.n71

Public Health England. (2020). Disparities in the risk and outcomes of COVID-19, 89. Retrieved from https://assets.publishing.service.gov.uk/government/uploads/system/uploads/attachment_data/file/892085/disparities_review.pdf%0Ahttps://www.gov.uk/government/publications/covid-19-review-of-disparities-in-risks-and-out

Public Health England. (2021a). COVID-19: Guidance for maintaining services within health and care settings Infection prevention and control recommendations Version 1.2, 1–49.

Public Health England. (2021b). Effectiveness of IPC measures in reducing COVID-19 transmission in adult and social care settings: rapid review protocol.

Rowe, T., Patel, M., O’Conor, R., McMackin, S., Hoak, V., & Lindquist, L. (2020). COVID-19 exposures and infection control among home care agencies. Archives of Gerontology and Geriatrics, 91(July), 104214. https://doi.org/10.1016/j.archger.2020.104214

Royal College of Nursing Wales. (2020). The Nursing Workforce in Wales 2020.

Royal College of Nursing Wales. (2021). NHS Pay Review Body Pay Round 2021-22. Royal College of Nursing (RCN Wales) evidence.

Sama, S. R., Quinn, M. M., Galligan, C. J., Karlsson, N. D., Gore, R. J., Kriebel, D., … Lindberg, J. E. (2021). Impacts of the COVID-19 Pandemic on Home Health and Home Care Agency Managers, Clients, and Aides: A Cross-Sectional Survey, March to June, 2020. Home Health Care Management and Practice, 33(2), 125–129. https://doi.org/10.1177/1084822320980415

Schweighart, R., Klemmt, M., Neuderth, S., & Teti, A. (2021). Experiences and perspectives of nursing home residents with depressive symptoms during the COVID-19 pandemic: a qualitative study. Zeitschrift Fur Gerontologie Und Geriatrie, 54(4), 353–358. https://doi.org/10.1007/s00391-021-01926-3

Shah, Z., Singh, V., Supehia, S., Mohan, L., Gupta, P. K., Sharma, M., & Sharma, S. (2021). Expectations of healthcare personnel from infection prevention and control services for preparedness of healthcare organisation in view of COVID-19 pandemic. Medical Journal Armed Forces India, 77, S459–S465. https://doi.org/10.1016/j.mjafi.2021.03.013

Sizoo, E. M., Monnier, A. A., Bloemen, M., Hertogh, C. M. P. M., & Smalbrugge, M. (2020). Dilemmas With Restrictive Visiting Policies in Dutch Nursing Homes During the COVID-19 Pandemic: A Qualitative Analysis of an Open-Ended Questionnaire With Elderly Care Physicians. Journal of the American Medical Directors Association, 21(12), 1774-1781.e2. https://doi.org/10.1016/j.jamda.2020.10.024

Social Care Institute for Excellence. (2021). Coronavirus (COVID-19) infection control for care providers. Retrieved from https://www.scie.org.uk/care-providers/coronavirus-covid-19/infection-control/quick-guide

Suárez-González, A., Rajagopalan, J., Livingston, G., & Alladi, S. (2021). The effect of Covid-19 isolation measures on the cognition and mental health of people living with dementia: a rapid systematic review of one year of evidence. MedRxiv. Retrieved from https://www.medrxiv.org/content/medrxiv/early/2021/03/20/2021.03.17.21253805.full.pdf

Telford, C. T., Bystrom, C., Fox, T., Holland, D. P., Wiggins-Benn, S., Mandani, A., … Shah, S. (2021). COVID-19 Infection Prevention and Control Adherence in Long-Term Care Facilities, Atlanta, Georgia. Journal of the American Geriatrics Society, 69(3), 581–586. https://doi.org/10.1111/jgs.17001

Telford, C. T., Onwubiko, U., Holland, D. P., Turner, K., Prieto, J., Smith, S., … Shah, S. (2020). Preventing COVID-19 Outbreaks in Long-Term Care Facilities Through Preemptive Testing of Residents and Staff Members — Fulton County, Georgia, March–May 2020. Morbidity and Mortality Weekly Report, 69(37), 1296–1299. https://doi.org/10.15585/mmwr.mm6937a4

The Joanna Briggs Institute. (2017). Checklist for Qualitative Research. The Joanna Briggs Institute, 6. Retrieved from http://www.joannabriggs.org/assets/docs/critical-appraisal-tools/JBI_Critical_Appraisal-Checklist_for_Qualitative_Research.pdf

Tulloch, J., Micocci, M., Buckle, P., Lawrenson, K., Kierkegaard, P., McLister, A., … Parvulescu, P. (2021). Enhanced Lateral Flow Testing Strategies in Care Homes Are Associated with Poor Adherence and Were Insufficient to Prevent COVID-19 Outbreaks: Results from a Mixed Methods Implementation Study. SSRN Electronic Journal, 1–8. https://doi.org/10.2139/ssrn.3822257

UK Government. (2021). Coronavirus guidance. Retrieved from https://www.gov.uk/guidance/covid-19-coronavirus-restrictions-what-you-can-and-cannot-do#wear-a-face-covering

Van der Roest, H. G., Prins, M., van der Velden, C., Steinmetz, S., Stolte, E., van Tilburg, T. G., & de Vries, D. H. (2020). The Impact of COVID-19 Measures on Well-Being of Older Long-Term Care Facility Residents in the Netherlands. Journal of the American Medical Directors Association, 21(11), 1569–1570. https://doi.org/10.1016/j.jamda.2020.09.007

Vardoulakis, S., Sheel, M., Lal, A., & Gray, D. (2020). COVID-19 environmental transmission and preventive public health measures. Australian and New Zealand Journal of Public Health, 44(5), 333–335. https://doi.org/10.1111/1753-6405.13033

Verbeek, H., Gerritsen, D. L., Backhaus, R., Boer, B., Koopmans, R., & Hamers, J. (2020). Allowing Visitors Back in the Nursing Home During the COVID-19 Crisis: A Dutch National Study Into First Experiences and Impact on Well-Being. Journal of the American Medical Directors Association, 21(7), 900–904. Retrieved from https://www.jamda.com/article/S1525-8610(20)30526-0/fulltext

Veritas Health Innovation. (2021). Covidence systematic review software. Melbourne, Australia.

Welsh Government. (2021). Care Homes Action Plan -Final update. Retrieved from https://gov.wales/care-homes-action-plan-final-update-html

Woolf, K., McManus, I. C., Martin, C. A., Nellums, L. B., Guyatt, A. L., Melbourne, C., … Pareek, M. (2021). Ethnic differences in SARS-CoV-2 vaccine hesitancy in United Kingdom healthcare workers: Results from the UK-REACH prospective nationwide cohort study. The Lancet Regional Health - Europe, 000, 100180. https://doi.org/10.1016/j.lanepe.2021.100180

